# Publication status and disclosure gaps in a cohort of 71 clinical trials assessing the safety and efficacy of 3 COVID-19 vaccines developed by Chinese biopharmaceutical companies: An observational cohort study

**DOI:** 10.1101/2024.03.17.24304386

**Authors:** Till Bruckner, Yixuan Chen, Carolina Cruz, Christie Ebube Dike, Belen Chavarria, Shiyu Chen, Ernest Dela Dzidzornu, Martin Ringsten

## Abstract

Transparency shortcomings can undermine confidence in the safety and efficacy of vaccines. This study assesses the publication status and disclosure gaps in a cohort of 71 clinical trials assessing the safety and efficacy of 3 COVID-19 vaccines developed by Chinese biopharmaceutical companies that received a World Health Organization Emergency Use Listing (EUL) and have been marketed globally. We searched trial registries and the scientific literature to assess the completion status of those 71 trials, and to determine whether the outcomes of completed trials have been made publicly available.

The 71 trials in our cohort were initiated by sponsors headquartered in 17 different countries and aimed to enrol a total of 313,750 participants from across 27 countries. Out of those 71 trials, 49 trials (69%) had verifiably been concluded. We were unable to determine the completion status of the remaining 22 trials (31%) with certainty. Depending on whether those 22 trials were assumed to have been concluded or not, we found that between 13 completed trials (27%) and 35 completed trials (49%) remained unreported. At least 9 trials (13% of the total) had not made their results public more than one year post completion. According to registry data, between 36,498 people (12% of participants across all trials) and 89,224 people (28%) had participated in trials that had been concluded and whose outcomes remained unreported. There were no tabular summary results available on trial registries for any of the trials.

Our findings suggest that there are significant gaps in clinical trial governance, especially in countries that have only recently emerged as significant players in biomedical research. Maintaining global progress in clinical trial reporting will require legislators and regulators to adopt and effectively enforce clinical trial reporting requirements that reflect WHO best practices. The global clinical trial registry infrastructure needs to be strengthened so that users can reliably determine whether studies have been withdrawn, are still ongoing, or have been concluded.

**Key messages:** *What is already known on this topic:* The results of many clinical trials are only made public partially, after long delays, or not at all. Such disclosure gaps can make it difficult to assess the benefits and harms of treatments, and can undermine public trust in health interventions, including vaccines. In North America and Europe, disclosure has significantly improved in recent years.

*What this study adds:* Previous research in this field has overwhelmingly focused on clinical trials run by sponsors in North America and Europe that usually involved patients in these regions. Our study provides a global perspective on the problem, using a cohort of trials with high salience for global public health. More than 1.3 billion doses of 3 COVID-19 vaccines originally developed in China have been exported to dozens of countries worldwide. We assessed the publication status and disclosure gaps of 71 clinical trials of these vaccines. We found widespread research activity in newly emerging hubs of biomedical innovation. In total, sponsors from 17 different countries and participants from 27 countries were involved in relevant trials. None of the concluded trials had reported outcomes in line with global best practice standards set out by the World Health Organisation.

*How this study might affect research, practice or policy:* Clinical trial activity is increasingly becoming global. Our research points to significant gaps in clinical trial governance in many emerging hubs of biomedical innovation. Legislators and regulators in these countries should ensure that all clinical trial results are rapidly made public in line with World Health Organisation best practices to prevent the growth of gaps in the global medical evidence base. In addition, there is an urgent need to strengthen the global clinical trial registry infrastructure.

## 1. Introduction

After the emergence of the SARS-CoV-2 virus in late 2019, the rapid development of vaccines became a global health priority, despite concerns that regulators might rush through approvals without due regard to vaccine safety and efficacy (1). Transparency in clinical trials of vaccines is essential to foster public trust and boost vaccine acceptance (2, 3).

However, the existing literature suggests that disclosure of the designs and outcomes of some COVID-19 vaccine trials has been suboptimal. A 2021 global report by Transparency International found that clinical trial protocols had been made public for only 12% of 86 clinical trials of 20 COVID-19 vaccines (4). Several Western pharmaceutical companies have been criticised for incomplete disclosure of trial designs (5). Vaccine approvals by the Indian drug regulator did not follow established pathways and decision-making processes were opaque (6). Chinese companies in particular have been criticised for only releasing partial trial results (7, 8).

As of August 2023, a total of 3 COVID-19 vaccines developed by 3 different Chinese companies had successfully completed the World Health Organisation’s (WHO’s) Emergency Use Listing (EUL) evaluation process, opening the door to international markets (9, 10). As of December 2022, more than 1.3 billion doses of these 3 vaccines had been exported to dozens of countries, significantly increasing the diversity and volume of global vaccine supply (11, 12).

**Table 1:** Key Chinese COVID-19 vaccine exports as of December 2022.

| Company | Vaccine | EUL date | Doses exported | Countries |
| --- | --- | --- | --- | --- |
| Sinopharm | Covilo | 07 May 2021 | 575.8 million | 93 |
| Sinovac | CoronaVac | 01 June 2021 | 774.2 million | 56 |
| CanSino | Convidecia | 19 May 2022 | 32.7 million | 10 |

Source: China COVID-19 Vaccine Tracker ([https://bridgebeijing.com/our-publications/our-publications-1/china-covid-19-vaccines-tracker/#Sinovac 8211 CoronaVac COVID-19 Vaccine](https://bridgebeijing.com/our-publications/our-publications-1/china-covid-19-vaccines-tracker/#Sinovac_8211_CoronaVac_COVID-19_Vaccine))

This paper examines a key element of trial transparency by assessing the publication status of all clinical trials involving the 3 COVID-19 vaccines developed by Chinese companies that received an EUL from the WHO. While Chinese law requires vaccine trial results to be shared with the national medicines regulator, it does not require them to be made public (13).

However, irrespective of national laws, publication of trial results is an ethical obligation under the Declaration of Helsinki (14). According to global best practices set out in the WHO’s Joint statement on public disclosure of results from clinical trials, the results of all clinical trials should be published on a public trial registry within 12 months of trial completion, and in a peer-reviewed journal within 24 months of trial completion (15). Sharing vaccine trial results with the public can support a better understanding of vaccine effectiveness and potential adverse effects (16-18).

The objective of this study was to assess whether the outcomes of clinical trials involving the 3 COVID-19 vaccines had been made public.

## 2. Methodology

The study protocol was preregistered on OSF (https://osf.io/4f9k7). No UK Health Research Authority NHS REC ethics approval was required as the study exclusively used publicly available study-level data. Patients and the public were not involved in this research. Outcomes are reported in line with the STROBE (STrengthening the Reporting of OBservational studies in Epidemiology) guideline for cohort studies (19). The study protocol and dataset are publicly available on OSF (https://osf.io/r3vp9/). This study did not receive external funding. The authors have no conflicts of interest to declare.

### 2.1 Cohort selection

We extracted the registry numbers of 95 clinical trials involving the 3 COVID-19 vaccines from McGill University’s COVID-19 Vaccine Tracker website (https://covid19.trackvaccines.org/), which had last been updated on 02 December 2022 (11). We de-duplicated 20 entries for 16 trials, creating a study cohort of 75 unique trials.

#### Data collection and search strategy

First, we entered the trial registry ID numbers sourced from the COVID-19 Vaccine Tracker into the ICTRP search function (https://trialsearch.who.int/Default.aspx) to identify potential additional registry entries for each of the 75 trials, and captured those.

Second, we manually extracted the following data from registry entries for each trial: (lead) sponsor, sponsor country, trial locations, trial phase, current trial status, trial completion date (primary if available, else overall), and number of trial participants (actual if available, else planned enrolment). If a trial was registered on multiple registries, we used registry data from registries in the following order of priority: ClinicalTrials.gov, ChiCTR, other registries. Registry data were first extracted in August 2023.

Third, two researchers independently searched for the outcomes of each of the 75 trials using a detailed search strategy set out in the study protocol (https://osf.io/4f9k7). Briefly, we (a) scanned the “results” section of all relevant trial registries for tabular summary results or hyperlinked publications, (b) searched PubMed and Google Scholar for the clinical trial ID number(s), (c) searched PubMed and Google Scholar for the trial title and principal investigator. The three steps above were completed between 23 August 2023 and 05 September 2023.

We then reviewed all journal publications to verify that they described the outcome of the trial by matching the 4 PICO criteria (patient inclusion criteria, intervention, comparison and outcomes). As per protocol, we only counted tabular summary results on trial registries and full-length publications of final trial results in peer-reviewed journals as ‘reported’. We classified publications presenting only interim trial outcomes, preprints, conference abstracts, press releases or other grey literature as ‘unreported’. For each trial, 2 team members, independently and in duplicate, reviewed the search results. Any inconsistencies and ambiguities were resolved by consensus or by a third team member. We then extracted the date of each publication.

Between 20 March 2024 and 13 April 2024, to provide additional quality assurance, we updated the spreadsheet to capture recent changes in registry entries, repeated the PICO match for previously located publications, and fully replicated the search for outcomes of trials for which no outcome had previously been located. We removed 4 trials that according to registry data had been withdrawn and never recruited participants (NCT05156632, NCT05254236, NCT05308602, NCT05442684), leaving a study cohort of 71 trials.

### 2.2 Outcome measures

The prespecified primary outcome measure was the number and percent of concluded (i.e. completed or terminated) trials whose results remained unreported. Secondary outcome measures were the number and percentage of participants enrolled (planned or actual enrolment, depending on registry data availability for each trial) in concluded trials whose results remained unreported, and the number and percentage of trials that met both WHO best practices in clinical trial reporting, i.e. results posted onto a public registry within 12 months of trial conclusion and results published in a peer-reviewed journal within 24 months of trial conclusion.

For the purpose of calculating the metrics above, as per our protocol, we considered trials to have been concluded if their status was marked as ‘completed’ or ‘terminated’ on a trial registry, and/or if the final outcomes had been made public on a trial registry or in a peer-reviewed journal (even if the registry still listed the trial as ongoing).

## 3. Results

### 3.1 Study population

According to registry data, the 71 trials in our cohort aimed to enroll a total of 313,750 participants from across 27 countries (Argentina, Bahrain, Brazil, Chile, China, Colombia, Egypt, Ghana, Guinea, Indonesia, Iran, Jordan, Laos, Madagascar, Malaysia, Mexico, Mozambique, Pakistan, Panama, Peru, Philippines, Russia, South Africa, Thailand, Turkey, United Arab Emirates, United States). The trials were initiated by a diverse set of commercial and non-commercial sponsors headquartered in a total of 17 countries (Argentina, Brazil, China, Chile, France, Indonesia, Iran, Pakistan, Peru, Philippines, Russia, Singapore, South Korea, Thailand, Turkey, United Arab Emirates, United States). The earliest and latest ‘actual’ trial (primary) completion dates listed in registries were in July 2020 and January 2024, respectively.

### 3.2 Trial status

According to registry data, 31/71 trials (44%) had been completed or terminated as of 13 April 2024. A trial is only listed as ‘completed’ or ‘terminated’ on a registry if the record holder has actively performed an update after the initial registration, so those trials had verifiably been concluded. The remaining 40/71 trials (46%) were not listed as having been concluded, including 12 trials whose status was listed as ‘unknown’.

**Table 3:** Trial status as per registry data, April 2024; n=71.

| **Trial status** | **Number of trials** | **Interpretation** |
| --- | --- | --- |
| Completed | 29 | Verifiably concluded as per registry data, n=31 |
| Terminated | 2 |  |
| Active not recruiting | 11 | Not verifiably concluded as per registry data, n=40 |
| Not yet recruiting | 7 |  |
| Recruiting | 9 |  |
| Suspended | 1 |  |
| Unknown | 12 |  |

### 3.3 Publication status

We located publications in peer-reviewed journals containing the final outcomes for 36/71 trials (51%); only 18 of those were listed as ‘completed’ or ‘terminated’ in registries. As per protocol, we did not classify 3 trials as reported as available publications only reported interim outcomes (NCT05323461, NCT04992260) or were only available as a preprint (NCT05137418). We only located outcomes uploaded onto registries for two trials (IRCT20140818018842N24, IRCT20201214049709N3); the outcomes of those trials had previously already been reported in peer-reviewed journals.

The outcomes of a further 13/71 (20%) verifiably concluded trials (11 completed and 2 terminated) had not been made public, including 9 trials that according to their ‘actual primary completion date’ field in ClinicalTrials.gov had concluded more than a year before our assessment.

### 3.4 Combining trial status and publication status

By combining trial status and publication status, we determined that 49/71 trials (69%) had definitely been concluded. Of these 49 concluded trials, 36/49 (73%) had published outcomes while 13/49 (27%) had not. Of the 13 unreported trials, 4 had been concluded less than a year ago; the remaining 9 had failed to meet the WHO benchmark of making outcomes public within one year of concluding the trial.

The remaining 22/71 trials (31%) were not listed as concluded in trial registries and had not made their outcomes public. However, the true status of these 22 trials was impossible to verify because trial status information in registries was often incorrect. For example, 18/36 trials with published outcomes (50%) in our cohort remained listed as not concluded in registries because their sponsors had failed to update their status after trial end. Thus, we categorised the status of the remaining 22 trials as ‘uncertain’.

**Table 4:**
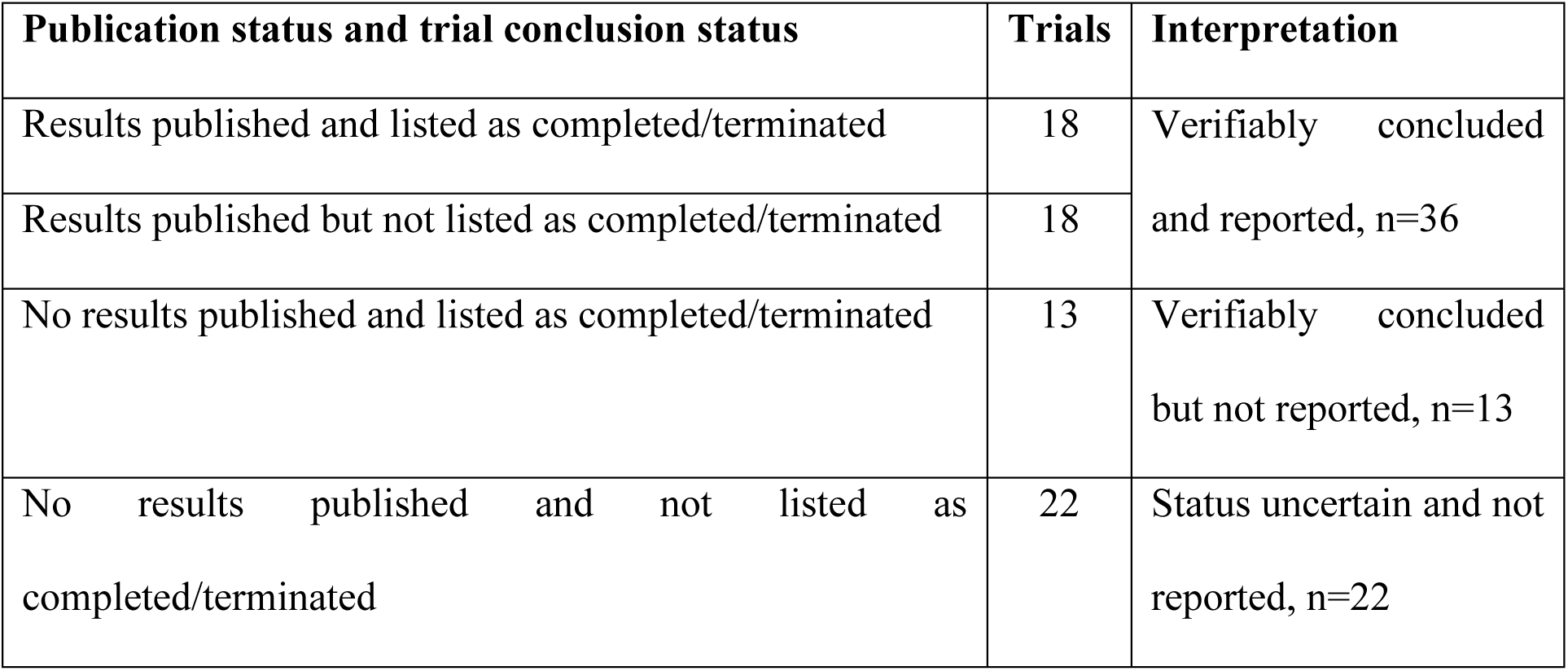
Outcome publication status and trial conclusion status on registries, April 2024; n=71.

| Publication status and trial conclusion status | Trials | Interpretation |
| --- | --- | --- |
| Results published and listed as completed/terminated | 18 | Verifiably concluded and reported, n=36 |
| Results published but not listed as completed/terminated | 18 |  |
| No results published and listed as completed/terminated | 13 | Verifiably concluded but not reported, n=13 |
| No results published and not listed as completed/terminated | 22 | Status uncertain and not reported, n=22 |

### 3.5 Primary outcome

The pre-specified primary outcome measure of this study was the number and percentage of concluded trials whose outcomes remained unreported. As we could not reliably determine whether 22 trials had concluded or not, we present our findings using two different baseline scenarios. The first scenario assumes that all 22 trials with uncertain status were still ongoing. The second scenario assumes that these 22 trials had all been concluded, i.e. completed or terminated. While we were unable to determine the precise number and percentage of concluded trials whose outcomes remained unreported, the actual figure lies within the range bracketed by these two scenarios.

**Table 5:** Number and percentage of concluded trials that remain unreported, April 2024.

|  | Concluded<br>trials (n) | Unreported<br>trials (n) | Unreported trials<br>(%) |
| --- | --- | --- | --- |
| Scenario 1: Verifiably concluded trials<br>only<br>(assumes that all 22 trials with uncertain<br>status are still ongoing) | 49 | 13 | 27% |
| Scenario 2: Verifiably concluded trials<br>plus uncertain trials<br>(assumes that all 22 trials with uncertain<br>status have been concluded) | 71 | 35 | 49% |

Thus, out of 71 clinical trials of 3 COVID-19 vaccines developed by Chinese companies that received a World Health Organization Emergency Use Listing (EUL) and have been marketed globally, a minimum of 13 trials (27%) and a maximum of 35 trials (49%) remained unreported as of 14 April 2024. Note that these figures include 4 recently concluded trials that had not yet exceeded the one-year results disclosure timeframe set out by the WHO and may yet report their results in a timely manner.

### 3.6 Secondary outcomes

This study had two pre-specified secondary outcome measures. The first was the number and percentage of participants in concluded trials whose outcomes remained unreported. Again, we present minimum and maximum figures using the two baseline scenarios described above, using the actual (or where not available, planned) enrolment figures as stated in trial registries.

**Table 6:** Number and percentage of participants in concluded unreported trials.

|  | Concluded<br>trials<br>(n) | Participants in<br>concluded trials<br>(n) | Participants in<br>unreported trials<br>(n) | Participants in<br>unreported trials<br>(%) |
| --- | --- | --- | --- | --- |
| Scenario 1: Verifiably<br>concluded trials only<br>(assumes that all 22<br>trials with uncertain<br>status are still ongoing) | 49 | 261,024 | 36,498 | 14% |
| Scenario 2: Verifiably<br>concluded plus<br>uncertain trials<br>(assumes that all 22<br>trials with uncertain<br>status have concluded) | 71 | 313,750 | 89,224 | 28% |

According to these calculations, out of the 313,750 people who participated in all 71 trials according to registry data, a minimum of 36,498 people (12%) and a maximum of 89,224 people (28%) had participated in trials that had been concluded and whose outcomes remained unreported as of 14 April 2024.

Our other secondary outcome measure was the number and percentage of trials that met both of two key WHO best practices in clinical trial reporting: posting outcomes onto an ICTRP-linked trial registry within 12 months of trial conclusion, and publishing outcomes in a peer-reviewed journal within 24 months of trial conclusion.

We expected to locate tabular summary results for some trials on the European EudraCT registry and/or on ClinicalTrials.gov. However, none of the trials in our cohort were registered on EudraCT, and none of the 62 trials registered on ClinicalTrials.gov had tabular summary results available there. We only located outcomes uploaded onto registries for 2 trials (IRCT20140818018842N24, IRCT20201214049709N3), both on the Iranian trial registry. In both cases, the responsible parties had copied and pasted a manuscript abstract into the results section of the registry following the manuscript’s publication in a peer-reviewed journal, rather than uploading tabular summary results. This approach improves the findability of trial outcomes. However, it fails to realise two other key advantages of registry reporting in tabular summary results format compared to journal publication: accelerated reporting timelines, and improved completeness and accuracy of outcome data (20-25). Thus, while between 2/49 (4%) of trials (Scenario 1) and 2/71 (3%) of trials (Scenario 2) technically met WHO best practices in clinical trial reporting, no trial fully delivered on the benefits of registry reporting.

## 4. Strengths and limitations

Our study has high relevance to public health and regulatory policy because it assesses the publication status and reporting gaps in clinical trials of 3 vaccines that have collectively been exported to dozens of countries worldwide and administered to hundreds of millions of people. However, it has several limitations. First, we relied on an external dataset generated by a different team of researchers to compile our initial study cohort of trials. Second, our follow-up period ranged from only 4 months to just under 4 years, making it likely that additional results will be published in future. Third, we may have missed relevant publications if these did not include the trial registry number and were in a language other than English. Finally, responsible parties had frequently failed to update trial registry entries as recommended by the WHO. This made it impossible to determine with certainty whether 22 of the trials in our cohort had been concluded. It also made it impossible to determine precisely how many people had participated in some concluded trials; we used planned enrolment figures where final enrolment numbers were not available.

## 5. Discussion

Our study shows that out of 71 clinical trials of 3 COVID-19 vaccines developed by Chinese companies that received a World Health Organization Emergency Use Listing (EUL) and have been marketed globally, a minimum of 13 trials (27%) and a maximum of 35 trials (49%) remained unreported post conclusion as of 14 April 2024, including at least 9 trials that had been concluded more than one year earlier. At first glance, the range of 27%-49% of trials remaining unreported appears to be broadly in line with non-reporting rates documented for large cohorts of clinical trials conducted in the United States and European Union (26-28). However, such direct comparisons would be misleading as those previous studies had longer follow-up periods and included a wide variety of trial types, including investigator-initiated studies of non-drug interventions that typically have below average publication rates.

Due to the lack of consistent updating of registry data, our finding that up to 89,224 people (28% of all trial participants) had been enrolled in trials that remained unreported should be treated with caution. The outcomes of only 2 trials (3% of all 71 trials) had been uploaded onto trial registries as recommended by the WHO.

Our findings suggest that there are significant gaps in clinical trial governance, especially in countries that have only recently emerged as significant players in biomedical research. Both commercial and non-commercial sponsors in the United States and many European countries have in recent years significantly improved their compliance with clinical trial disclosure requirements (29, 30). Legislation in both the United States and the European Union requires sponsors to disclose the outcomes of many interventional studies, including vaccine trials, in the form of tabular summary results uploaded onto a trial registry within a year of concluding a trial (31-33). The WHO also recommends this reporting format and timeframe.

In contrast, we found that the final outcomes for 9 COVID-19 vaccine trials remained unreported more than one year after the end of the trial. All of these trials had recruited participants exclusively in emerging hubs of biomedical innovation: Argentina, China, Chile, Peru, the Philippines, and the United Arab Emirates. Either these jurisdictions do not have adequate disclosure laws, or they do not effectively enforce legal compliance, or both. Similarly, none of the 36 reported trials in our study had reported outcomes on a registry using the tabulated summary results format recommended by the WHO; one of them (NCT05293665, sponsored by the American company Vaxxinity) appeared to be in violation of United States disclosure laws (34). Maintaining global progress in clinical trial reporting will require legislators and regulators worldwide, and in particular in emerging innovation hubs, to adopt and effectively enforce clinical trial reporting requirements that reflect WHO best practices.

Finally, our research highlights the urgent need to strengthen and adequately fund the global clinical trial registry infrastructure. The WHO-run International Clinical Trials Registry Platform (ICTRP) currently aggregates data from a network of 19 individual registries. Largely due to widespread data quality issues within individual registries, this infrastructure failed to provide a comprehensive, accurate and up-to-date overview of the global medical research landscape during the early months of the COVID-19 pandemic. This led to the proliferation of redundant studies and research waste on a large scale (35). Our study documents that these data quality issues persist. We were unable to reliably determine how many trials of 3 widely administered vaccines were still ongoing, how many had been concluded, how many people had participated in them, and what harms and efficacies had been detected. Notably, we were completely unable to access the Peruvian trial registry’s online platform during both assessment phases of this study. We were also unable to determine which countries two trials (NCT04984408, NCT05308576) were being conducted in. While there is no substitute for adequate funding, the Thai registry’s recent introduction of automated email reminders (36) and the ISRCTN registry’s forthcoming transparency tracker (37) illustrate that in some cases, significant improvements can be achieved at minimal cost.

**Table 7:** List of all 71 vaccine trials included in this study.

| **Trial ID number(s)** | **Phase** | **Enrolment** | **Verifiably concluded?** | **Results public?** |
| --- | --- | --- | --- | --- |
| NCT05374954 | 3 | 4200 | No | Unreported |
| NCT04998240 | 2 | 360 | No | Unreported |
| NCT05226429 | 1 & 2 | 495 | No | Unreported |
| NCT05463419<br>PHRR230223-005230 | 2 & 3 | 5860 | No | Unreported |
| NCT05005156 | 2 | 876 | No | Unreported |
| NCT05225285 | 3 | 1120 | No | Unreported |
| NCT05163652 | 3 | 432 | No | Unreported |
| NCT05433272 | 3 | 1400 | No | Unreported |
| PACTR202204735722575 | 3 | 10000 | No | Unreported |
| NCT05409300 | 2 | 200 | No | Unreported |
| NCT05172193 | 2 | 600 | No | Unreported |
| IRCT20171122037571N3 | 1 & 2 | 500 | No | Unreported |
| NCT05465902 | 2 | 600 | No | Unreported |
| NCT04984408 | 3 | 8825 | No | Unreported |
| NCT05308576 | 3 | 10000 | No | Unreported |
| NCT05162482 | 2 | 1680 | No | Unreported |
| NCT05428592 | 3 | 1100 | No | Unreported |
| NCT05293665 | 3 | 944 | No | Unreported |
| NCT05463354 | 2 | 450 | No | Unreported |
| NCT05150496 | 2 | 640 | No | Unreported |
| NCT05230940 | 2 | 644 | No | Unreported |
| NCT05323461 | 3 | 1800 | No | Unreported |
| NCT04352608 | 1 & 2 | 744 | Yes | Reported |
| NCT04341389<br>ChiCTR2000031781 | 2 | 508 | Yes | Reported |
| NCT04456595 | 3 | 12688 | Yes | Reported |
| NCT04508075<br>INA-WXFM0YX | 3 | 1620 | Yes | Reported |
| NCT04313127<br>ChiCTR2000030906 | 1 | 108 | Yes | Reported |
| NCT04551547 | 1 & 2 | 552 | Yes | Reported |
| NCT04510207<br>ChiCTR2000034780 | 3 | 44101 | Yes | Reported |
| NCT04540419 | 3 | 500 | Yes | Reported |
| NCT04962906 | 2 | 192 | Yes | Reported |
| NCT04526990 | 3 | 44247 | Yes | Reported |
| NCT04566770 | 2 | 480 | Yes | Reported |
| NCT04383574 | 1 & 2 | 422 | Yes | Reported |
| NCT04979949 | 2 | 222 | Yes | Reported |
| NCT05204589 | 3 | 10420 | Yes | Reported |
| NCT04582344 | 3 | 10214 | Yes | Reported |
| NCT04840992 | 1 & 2 | 840 | Yes | Reported |
| NCT04651790 | 3 | 2300 | Yes | Reported |
| NCT05583357 | 1 | 40 | Yes | Reported |
| IRCT20140818018842N24 | 2 & 3 | 61 | Yes | Reported |
| NCT04942405 | 3 | 1290 | Yes | Reported |
| NCT04983537 | 2 | 120 | Yes | Reported |
| NCT04988048 | 2 | 1760 | Yes | Reported |
| NCT05087368 | 2 | 520 | Yes | Reported |
| NCT04992182 | 2 | 534 | Yes | Reported |
| NCT04800133 | 3 | 1018 | Yes | Reported |
| NCT05043259 | 1 & 2 | 420 | Yes | Reported |
| ChiCTR2000032459 | 1 & 2 | 640 | Yes | Reported |
| IRCT20210206050259N3 | 3 | 41128 | Yes | Reported |
| NCT05109559 | 1 & 2 | 690 | Yes | Reported |
| TCTR20210720005 | 2 | 180 | Yes | Reported |
| TCTR20210920005 | 2 | 500 | Yes | Reported |
| NCT04568811 | 1 | 89 | Yes | Reported |
| NCT05049226 | 2 | 1250 | Yes | Reported |
| NCT05249816 | 3 | 1000 | Yes | Reported |
| IRCT20201214049709N3 | 3 | 41128 | Yes | Reported |
| NCT05580159 | 3 | 2000 | Yes | Reported |
| NCT04617483 | 3 | 1040 | Yes | Unreported |
| NCT04612972 | 3 | 12000 | Yes | Unreported |
| NCT04884685 | 2 | 500 | Yes | Unreported |
| NCT04560881;<br>BIBP2020003AR | 3 | 3000 | Yes | Unreported |
| NCT04917523 | 3 | 1800 | Yes | Unreported |
| NCT05323435 | 2 & 3 | 300 | Yes | Unreported |
| NCT05169008 | 3 | 91 | Yes | Unreported |
| NCT05137418 | 3 | 1000 | Yes | Unreported |
| NCT05077176 | 3 | 4340 | Yes | Unreported |
| NCT04992260 | 3 | 11349 | Yes | Unreported |
| NCT05228613 | 1 | 175 | Yes | Unreported |
| NCT05593042 | 2 | 551 | Yes | Unreported |
| PHRR210210-003308 | 2 & 3 | 352 | Yes | Unreported |

*Note: Full dataset publicly available on OSF: https://osf.io/r3vp9/*

## Data Availability

Key data produced in the present work are contained in the manuscript (Table 7). All data produced are available online on OSF: https://osf.io/r3vp9/

https://osf.io/4f9k7

## Notes

### Competing Interest Statement

The authors have declared no competing interest.

### Clinical Protocols

https://osf.io/4f9k7

### Funding Statement

This study did not receive any funding.

### Summary of Updates

Clinical trials were de-duplicated, additional searches for publications were run, sponsor names and countries were added, and the entire dataset was updated. These updates led to changes in the conclusions section.

